# Implementation of Mercury Biomonitoring in German Adults using Dried Blood Spot Sampling in Combination with Direct Mercury Analysis

**DOI:** 10.1101/2021.02.17.21251759

**Authors:** Ann-Kathrin Schweizer, Michael Kabesch, Caroline Quartucci, Stephan Bose-O’Reilly, Stefan Rakete

## Abstract

Venous blood is a preferred matrix for the determination of total mercury (Hg) in human biomonitoring but has some drawbacks such as the requirement for an uninterrupted cold chain for transport and storage and the need of medical personnel for sample collection. Therefore, we tested and implemented a simpler and less expensive method for measuring Hg in human blood using dried blood spots (DBS). For method development, we investigated the influence of different storage conditions (temperature, storage vessel, time) on DBS samples. For method validation, we compared DBS and venous blood and investigated whether DBS sampling is suitable for measuring Hg in the general population in countries with low Hg exposure such as Germany. Based on our results, we found that pre-cleaned glass tubes were most suitable for storage of DBS samples, as this allowed the samples to remain stable for at least four weeks even at high temperatures (40°C). When comparing venous blood and DBS, a very good correlation (r=0.95, p<0.01) and high precision of DBS (mean relative standard deviation 8.2% vs. 7.2% in venous blood samples) were observed. Comparing the recoveries of both matrices in different concentration ranges, the scattering of the recoveries decreases with increasing Hg concentration. The same applies to the mean recoveries. Overall, we found comparable results for DBS and whole blood using direct Hg analysis. Furthermore, we demonstrated that DBS are suitable for Hg biomonitoring in the general population in Germany and improved the storage conditions for the DBS.

## INTRODUCTION

Mercury (Hg) is a toxic metal and a hazard for humans and the environment.^1^ Even the low exposure to Hg has negative effects on the health of humans, e.g. on cardiovascular, reproductive, renal and central nervous systems; especially during pregnancy and infanthood the cognitive development can be disturbed at low levels of exposure.^2-4^ To be able to study these associations in detail, simple methods to sample Hg in those populations, e.g. newborns are needed. It can occur as elemental Hg, inorganic Hg (e.g. HgCl_2_) and organic Hg (e.g. methylmercury), each species having different toxicodynamic properties.^3,5^ The major anthropogenic sources of Hg emissions are the burning of fossil fuels and artisanal and small-scale gold mining (ASGM) activities.^6^ ASGM also poses a strong source of exposure to Hg, as Hg is used as a binding agent for gold and is mainly inhaled by miners in this process.^7-10^ ASGM areas are usually remote areas, and only with simplified methods, Hg in human specimens could be analyzed. In high-income countries, the main source of Hg exposure is the consumption of saltwater fish, e.g. tuna, which contains a high amount of Hg due to the accumulation in the food chain.^3,10^ The exposure due to fish consumption is associated with negative effects on the pre- and postnatal development.^3^

Screening of newborns for mercury levels is not yet performed regularly, e.g. in regions with high consumption of predatory fish. To assess the Hg exposure in human blood, venous blood represents the current gold standard. Challenges in using venous blood as a sample material include the necessary availability of trained medical personnel and the maintenance of an uninterrupted cooling chain prior to laboratory analysis. This may lead to relatively high logistical efforts and costs. Furthermore, venipuncture is a more invasive technique compared to other methods of specimen collection and is ethical concerning for certain populations such as infants and children.^8,9,11-15^ As an alternative to venous blood sampling, dried blood spot (DBS) sampling can be used for Hg biomonitoring, too.^8,9,11,13,15-19^ Here, capillary blood, e.g. from the finger, is collected on special filter cards. In contrast to venous blood sampling, only a few drops of blood (approximately 60µl per circle) is needed. DBS sampling usually do not require cooling, extensive storage space or even medically-trained personnel, therefore increasing the feasibility of this method for studies of large populations or in remote areas with limited laboratory infrastructure.^11,14-16,18,20,21^ So far, only three studies compared DBS with venous blood sampling for human biomonitoring of Hg.^9,13,15^ One study found an relationship between whole blood and DBS, but it was confounded by the high background contamination of DBS cards, which is why they considered the retrospective use of DBS without precleaning of the cards inappropriate.^13^ A second study showed that the Hg levels in DBS samples from pregnant women, who live in an ASGM area, were comparable to venous blood levels.^9^ Furthermore, it was demonstrated that DBS are comparable to associated whole blood levels with samples from low exposed persons, too, when methylmercury and inorganic mercury were analyzed using gas chromatography-cold vapor atomic fluorescence (GC-CVAFS).^15^ However, DBS sampling has so far not been used for biomonitoring of total Hg in blood in the general population in countries with an expected low exposure, e.g. Germany, in combination with direct Hg analysis. The implementation of DBS sampling for Hg biomonitoring comes with multiple challenges such as the background contamination of DBS cards as well as potential contamination or loss of Hg during sampling, transport and storage.^8,9,11,13,15-17,20^ Only few studies dealt with the storage stability of Hg in DBS samples.^15,16,19^ In these studies, Hg in DBS samples was found to generally stable under the investigated conditions. However, no studies have been carried out the influence of different storage vessels for DBS, although its storage is an important factor for possible contamination of DBS samples.

The aim of this study was the development and validation of a biomonitoring method for total Hg in capillary blood using DBS sampling in combination with direct Hg analysis. Therefore, DBS and venous blood samples were collected from German adults to evaluate, if both sampling methods will provide comparable results. Additionally, the influence of different storage vessels on the stability of Hg in DBS samples was investigated.

## MATERIAL AND METHODS

### Used Materials

Hg ICP standard (1g/l in 10 % nitric acid), hydrochloric acid (30 %) for trace metal analysis and nitric acid (65 %) for trace metal analysis were obtained from Merck (Darmstadt, Germany). Whatman^®^ 903 protein saver cards for DBS sampling were obtained from Sigma-Aldrich (St. Louis, USA). Disposable lancets (Solofix^®^) from B. Braun (Melsungen, Germany) were used for fingerpricks. LDPE plastic zip lock bags (22 × 16 cm) for DBS storage were obtained from Buerkle (Bad Bellingen, Germany). Glass tubes with plastic screw caps (98 × 16 mm) for DBS storage were obtained from Schuett-biotec (Goettingen, Germany). Certified reference materials for blood (ClinChek^®^) were obtained from RECIPE (Munich, Germany).

### Storage Stability of Hg in DBS

In order to investigate the influence of different storage conditions on Hg levels in DBS, fresh venous blood from one volunteer was used for the following experiments. In detail, three spots of one DBS card were spiked with 55 µl blood in order to be within the circled line and dried for two hours at room temperature. To test the influence of the storage vessel, the prepared DBS cards were individually stored in plastic bags, untreated glass tubes and glass tubes that had been pre-cleaned with an aqueous mixture of hydrochloric and nitric acid (each 5 %, v/v), respectively (Figure S1). In detail, the glass tubes were filled with 5 ml acid mixture and put on a roll mixer for two hours. Afterwards, the tubes were rinsed twice with ultrapure water and dried at 100 °C in an oven. In order to store DBS samples in the glass tubes, the DBS spots were cut out approximately 5 mm below the circled line using a stainless steel scissor. To test the influence of the storage temperature, each storage vessel was stored at −20 °C, room temperature and 40 °C for one, two and four weeks, respectively. Every single experiment was conducted in duplicate. As a reference, Hg levels were additionally analyzed in DBS cards prepared in the same way as described above immediately after drying. Furthermore, the stability of Hg in DBS was also tested using certified reference material for blood, storing the prepared DBS cards in plastic bags at 40 °C for up to four weeks.

### Application of DBS for Human Biomonitoring of Hg

#### Study design

This proof-of-principle study was conducted at the Institute and Clinic for Occupational, Social and Environmental Medicine, LMU University Hospital Munich. The study was carried out in accordance with the Code of Ethics of the Declaration of Helsinki for experiments involving human subjects and reviewed and approved by the ethics committee of the Ludwig Maximilians University of Munich (20-091). Participants had to be at least 18 years old and no individual restrictions for venous and capillary blood sampling. Prior to the sampling, each participant was informed about the study and signed an informed consent form. Each participant was asked to fill out a questionnaire about potential Hg exposure (e.g. fish consumption and dental amalgam).

#### Sample collection

Venous blood samples from all participants were collected into 7 ml Lithium-Heparin-coated tubes for trace metal analyses (Sarstedt^®^) and stored at −20 °C until analysis. For DBS sampling, the same participant was asked to wash his hands thoroughly to prevent contamination during the sampling. Afterwards, one finger was disinfected and pricked with a sterile disposable lancet. The first drop of blood was discarded before filling three spots of one DBS card with capillary blood. If the blood flow stopped before the circled area of three spots was completely filled, another finger was punctured with the consent of the participant. The DBS samples were dried for two hours at room temperature. To test the influence of storage on real samples, some DBS samples (n=18) were analyzed immediately after drying. The remaining samples (n=32) were stored for one week at room temperature in a pre-cleaned glass tube as described above.

### Sample Analysis

All samples were analyzed by direct Hg analysis using a DMA80-evo^®^ instrument (MLS Mikrowellen-Labor-Systeme GmbH, Leutkirch, Germany). Hg was detected by atomic absorption at 253.5 nm. The quantification was based on an external calibration. Before every analysis series, the sample boats were preconditioned to avoid interference by residual Hg. For venous blood, 100µl blood were directly pipetted into the sample boats and analyzed. Each venous blood sample was at least analyzed in triplicate. For DBS, three completely filled circles were punched out using a precleaned 0.5-inch stainless steel paper puncher. The punched circles, which contained approximately 60 µl (specification by the manufacturer, 55µl for stability experiments) blood, were individually placed directly into the sample boat and analyzed. Blanks were prepared in the same manner using an empty circle from the same DBS card. The limits of detection (LOD) were 0.02 µg/l for venous blood and 0.09 µg/l for DBS samples, respectively. The limits of quantitation (LOQ) were 0.04 µg/l for venous blood and 0.23 µg/l for DBS samples, respectively. For quality assurance, an aqueous Hg standard solution and certified reference material for whole blood (ClinCheck^®^, Recipe, Munich, Germany) were analyzed daily prior to the analysis of the samples.

### Statistical Analysis

For data processing, Excel 2016 was used. Statistical analysis was performed with IBM SPSS® Statistics, Version 25. Samples below the LOD or the LOQ were excluded from the statistical analysis. For evaluation of the accuracy of DBS, the recovery was calculated from the Hg levels in DBS and venous blood (Hg_DBS_/Hg_VB_*100 %). Differences in recoveries due to the storage conditions were analyzed using single factor ANOVA and post-hoc tests (Sheffé, Bonferoni). For the study with human subjects, descriptive statistics were used for information about the study population and Hg levels in venous blood and DBS samples. Correlation between Hg levels in DBS and venous blood were analyzed by Spearmen-Rho test. Jonksheere-Tepstra tests were performed to evaluate the differences in the recoveries between individual concentration ranges (Hg levels in venous blood: < 0.5 µg/l, 0.5 – 1.0 µg/l, 1.0 – 1.5 µg/l, > 1.5 µg/l). Data was graphically displayed using bar charts, scatter plots and Bland-Altman plot.

## RESULTS AND DISCUSSION

### Storage Stability of Hg in DBS

To evaluate the effect of different storage conditions, DBS samples were stored in different vessels at different temperatures and for different times. The mean Hg level in DBS samples when analyzed directly after drying was 0.81 µg/l (100 %). In **Figure 1**, the recoveries of Hg in DBS cards that have been stored under different conditions are shown. For pre-cleaned glass tubes (a), recoveries varied from 78 to 99 % and the differences between the groups were not significant. For plastic bags, recoveries varied between 70 and 240 %. For storage at −20 °C and room temperature, no significant differences between the groups were found. In contrast, recoveries significantly increased after two and four weeks of storage when the samples were kept at 40 °C (p<0.01). Hg recoveries in the uncleaned glass tubes showed high variability that could not be explained by the storage conditions (Figure S1). However, recoveries were mainly around 100 % for all temperatures.

**Figure 1:**
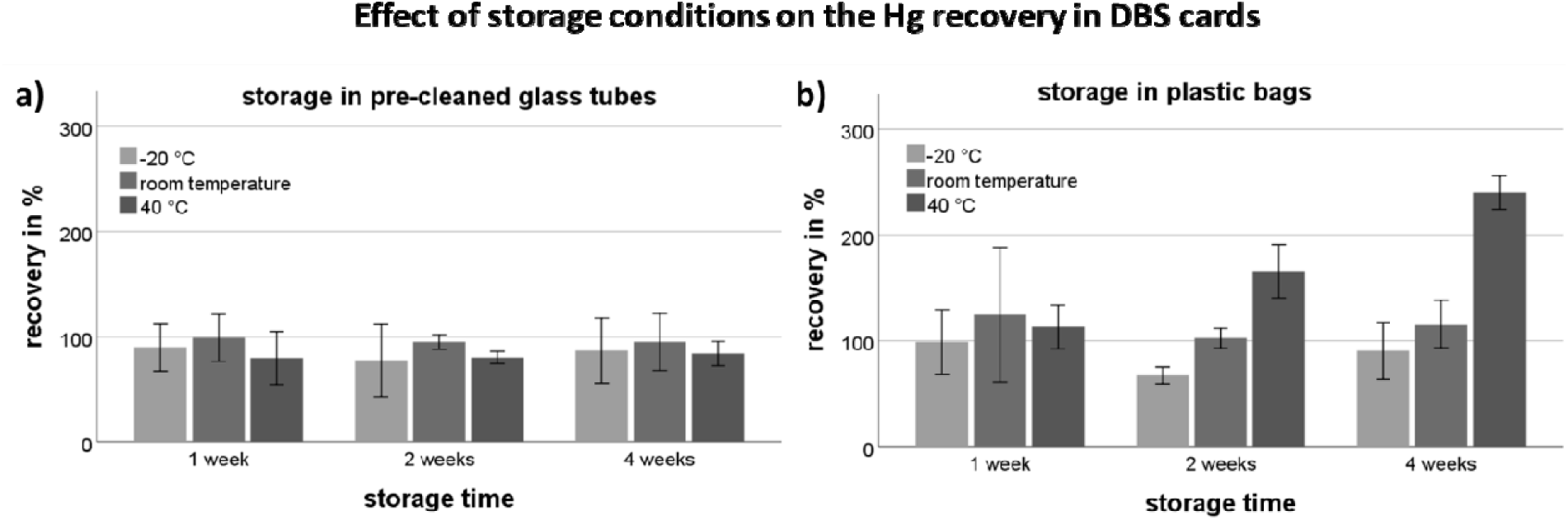
Effect of storage conditions (temperature, time) on the Hg recovery in DBS cards stored in pre-cleaned glass tubes (a) or plastic bags (b). Each bar shows the mean recovery from six individual DBS spots and the standard deviation.

Based on these results, we decided to use pre-cleaned glass tubes for the storage of DBS samples in the further course of this study. Consequently, we believe that the samples will be stable for at least four weeks, even when stored at 40 °C. Uncleaned glass showed good results, too. However, the results suggest that individual glass tubes may be contaminated with residual amounts of Hg, e.g. during manufacturing, and therefore require cleaning before they can be used as containers for DBS samples. The majority of previously published studies that use DBS samples for Hg biomonitoring use plastic bags for storage.^13,15,16,18,19^ Although the storage of DBS samples in plastic bags showed satisfying recoveries at −20 ° and room temperature in our experiments, Hg levels were significantly elevated when the samples were stored at 40 °C. The elevated Hg levels may be explained by residual Hg in the bags or ambient Hg that could penetrate the plastic bag and bind to the dried blood. Other studies recommend the use of metal-free plastic bags or cleaning procedures.^9,13^ However, we preferred glass tubes to plastic bags, as the handling for cleaning and drying of the tubes was relatively simple.

Interestingly, increased Hg levels in DBS samples were not observed when certified reference material for blood, which also has been used in several other studies was used instead of fresh blood for stability experiments (Figure S2).^8,9,11,13,15,16,18^ Furthermore, blank values were not affected by any storage condition, too (data not shown). Therefore, we assume that external Hg in any form only binds to DBS samples from fresh blood. Consequently, fresh blood should be preferably used for method development as it apparently has different properties compared to certified reference material.

### Correlation of Hg in Venous Blood and DBS Samples

In total, 53 participants were recruited. One participant was excluded because only one circle on the DBS card was completely filled. Two participants were excluded because of po sible sample contamination. In contrast, six participants with only two DBS spots used for Hg analysis were included in the study. Consequently, the data from 50 participants was used for further analysis. The results for venous blood and DBS samples are shown in **Table 1**. The Hg levels in one venous blood sample was below LOD, in another below the LOQ.

**Table 1:** Hg levels in venous blood and DBS samples of the study participants (n=50).

|  | GM<br>[μg/l] | Median<br>[μg/l] | Min.<br>[μg/l] | Max.<br>[μg/l] | Mean RSD*<br>[%] | samples < 20 % RSD*<br>n (%) |
| --- | --- | --- | --- | --- | --- | --- |
| **Venous blood** | 0.65 | 0.87 | < LOD | 4.35 | 7.2 | 45 (94) |
| **DBS** | 0.67 | 0.73 | < LOQ | 3.18 | 8.2 | 41 (93) |

GM: geometric mean, RSD: relative standard deviation from three individual analysis, LOQ: limit of quantitation, DBS dried blood spots, \* samples below the LOD/LOQ were excluded

For DBS samples, six samples were below the LOQ and therefore excluded from further analysis. Stratification of the Hg levels in venous blood by gender, age, fish consumption and dental amalgam fillings can be found in the Supporting Information (Table S1). The individual mean Hg values in venous blood and DBS samples can be found in the Supporting Information (Table S2).

Regarding the precision of Hg analysis, venous blood sampling was slightly but not significantly superior to DBS sampling. Although more than 50 % samples of both venous blood and DBS showed a relative standard deviation (RSD) of less than 10 %, mean RSD of venous blood samples was lower compared to DBS (7.2 % vs 8.2 %). Furthermore, 94 % of the venous blood samples were below an RSD of 20 % compared to 93 % of the DBS samples. This may be explained by residual Hg in the filter paper. As other studies report, residual Hg is likely not homogenously distributed in the filter paper, limiting the correction for blank samples from the same DBS card.^13^ Nevertheless, precision of DBS sampling was relatively comparable to venous blood sampling.

The correlation of the Hg levels in venous blood and DBS samples is shown in **Figure 2**. We found a very strong linear correlation between Hg levels in venous blood and DBS samples (r=0.95, p<0.001, Spearman-Rho). This is comparable to what has been found by Nyanza et al. and Santa-Rios et al.^9,15^ The majority of the samples were within the desired recovery range between 70 and 130 % (75 %). Overall, recoveries ranged between 62 and 210%. The associated Bland-Altman plot (**Figure 3**) further confirms that DBS sampling and venous blood sampling are comparable methods. However, the recovery of Hg levels in DBS samples depended on the concentration of Hg in venous blood.

**Figure 2:**
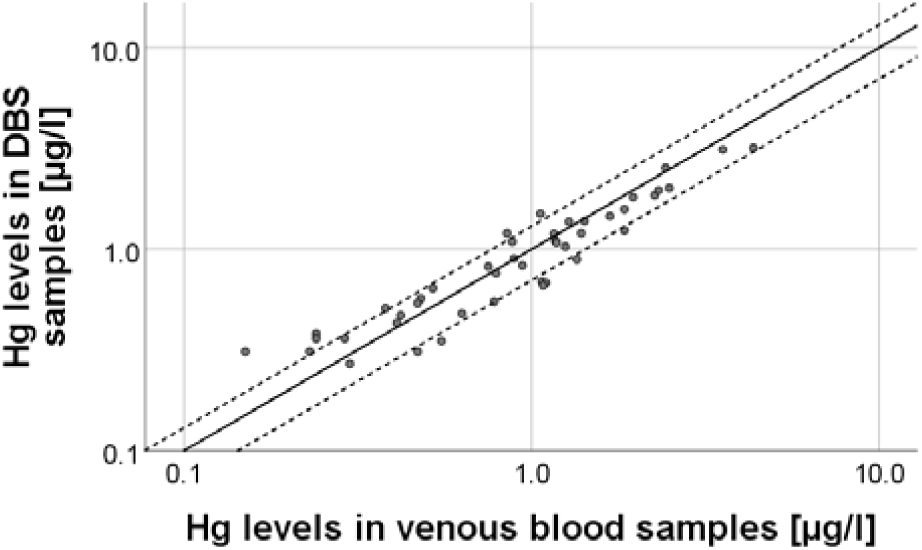
Correlation of Hg levels in venous blood and DBS samples (n=44). The straight line represents the identity line; the dashed lines delimit the desired DBS recovery range of 70 to 130 %.

**Figure 3:**
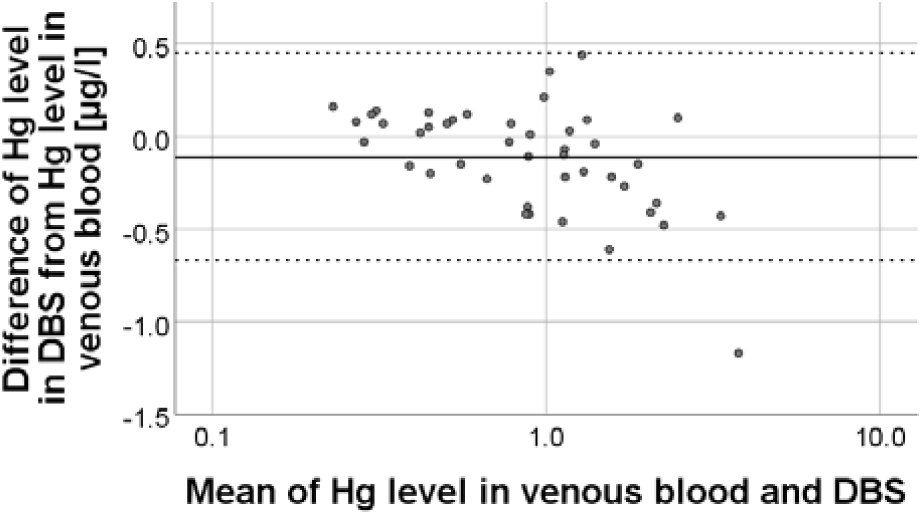
Bland-Altman plot of the absolute differences between Hg levels in venous blood and DBS samples vs the mean of Hg levels in both samples.

In **Figure 4**, the recoveries of DBS samples stratified by concentration ranges are shown. Median recoveries significantly decreased with increasing Hg levels in venous blood (p=0.001, Jonksheere Tepstra test). Additionally, scattering of the recovery was lowest DBS samples when Hg levels in venous blood were above 1.5 µg/l. This may be explained by the fact that residual Hg on the DBS cards have a higher impact on the results when the Hg levels in blood are relatively low.

**Figure 4:**
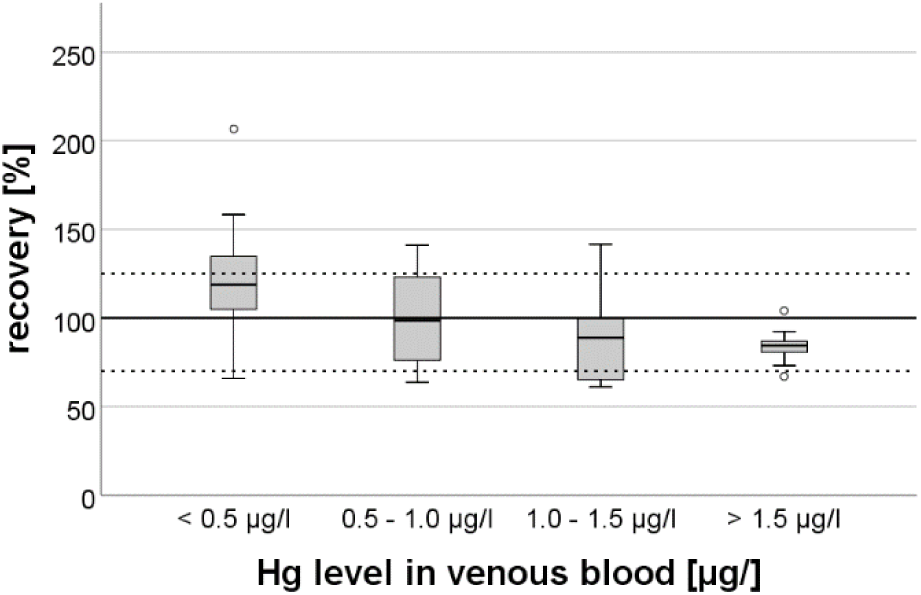
Box plot of the recoveries of Hg in DBS samples with different ranges of Hg levels in venous blood samples. The dotted lines resemble a recovery of 70 % and 130%, respectively.

Furthermore, it appears that results from the DBS samples tend to be elevated in samples with venous blood Hg levels below 0.5 µg/l. Above 0.5 µg/l, mean recoveries were found to be between 84 and 99 %. Because the impact of residual Hg decreases with increasing Hg levels in venous blood, lowest scattering of recoveries was found in samples above Hg levels of 1.5 µg/l. The fact that the recoveries for DBS samples with Hg levels above 1.0 µg/l were below 100 % could be due to a lower blood volume in the punched DBS than the estimated 60 µl. Other studies report the influence of hematocrit on the volume of blood in a specific area of DBS card and recommend the weighing of punched spots to adjust for individual hematocrit values.^9,22,23^ However, we deliberately did not weigh the punched DBS spots because the hematocrit levels in humans are in sufficiently narrow physiological ranges for meaningful results.^21^ Finally, no significant difference for Hg recoveries was found between direct analysis of the DBS cards after drying (n = 17; median recovery: 103 %) and analysis after storage for one week (n = 27; median recovery: 88 %). Consequently, the storage of DBS samples in pre-cleaned glass tubes successfully prevented contamination.

## STRENGTHS AND LIMITATIONS OF THE STUDY

The application of DBS sampling in combination with direct Hg analysis for Hg biomonitoring is the key strength of our study. Furthermore, our study sheds more light on the stability of Hg in DBS samples under different storage conditions, especially with regard to storage vessels and temperatures. Our study also emphasizes the importance of using of fresh blood samples for method development instead of certified reference materials.

A limitation of our study is the fact that the exact blood volume of each DBS was not adjusted for hematocrit levels as suggested in other studies.^9,11,13^ Nevertheless, Funk et al. state that estimated blood volume sufficient for valid results.^17^ Furthermore, the LOQ was too low for about 10 % of the DBS samples. This is due to the relatively low blood volume of 60 µl on one hand and varying residual Hg amounts on the DBS cards that interfere greatly with low Hg amounts from in the sampled blood on the other hand. Nonetheless, the LOQ of our method (0.24 µg/l), is, in our opinion, more than sensitive enough to identify people with elevated Hg levels in blood. However, a better sensitivity may be achieved by using precleaned DBS cards and ICP-MS for analysis as it has been described in other studies.^8,9,13,16-18^

## Data Availability

The data is available upon reasonable request

## ASSOCIATED CONTENT

### Supporting Information

The Supporting Information is available free of charge on the ACS Publications website.

Additional information of the participants and storage conditions (PDF).

## AUTHOR INFORMATION

### Author Contributions

**Ann-Kathrin Schweizer:** Sampling, sample analysis, data analysis, visualization, preparation of the original draft, writing - review & editing. **Michael Kabesch:** supervision, conceptualization, writing - review & editing. **Caroline Quartucci:** Sampling, supervision, writing - review & editing. **Stephan Bose-O’Reilly:** supervision, conceptualization, methodology, data analysis, writing - review & editing. **Stefan Rakete:** Conceptualization, methodology, funding acquisition, data analysis, writing - review & editing, preparation of the final manuscript. All authors have given approval to the final version of the manuscript.

Authors are required to submit a graphic entry for the Table of Contents (TOC) that, in conjunction with the manuscript title, should give the reader a representative idea of one of the following: A key structure, reaction, equation, concept, or theorem, etc., that is discussed in the manuscript. Consult the journal’s Instructions for Authors for TOC graphic specifications.

## Insert Table of Contents artwork here

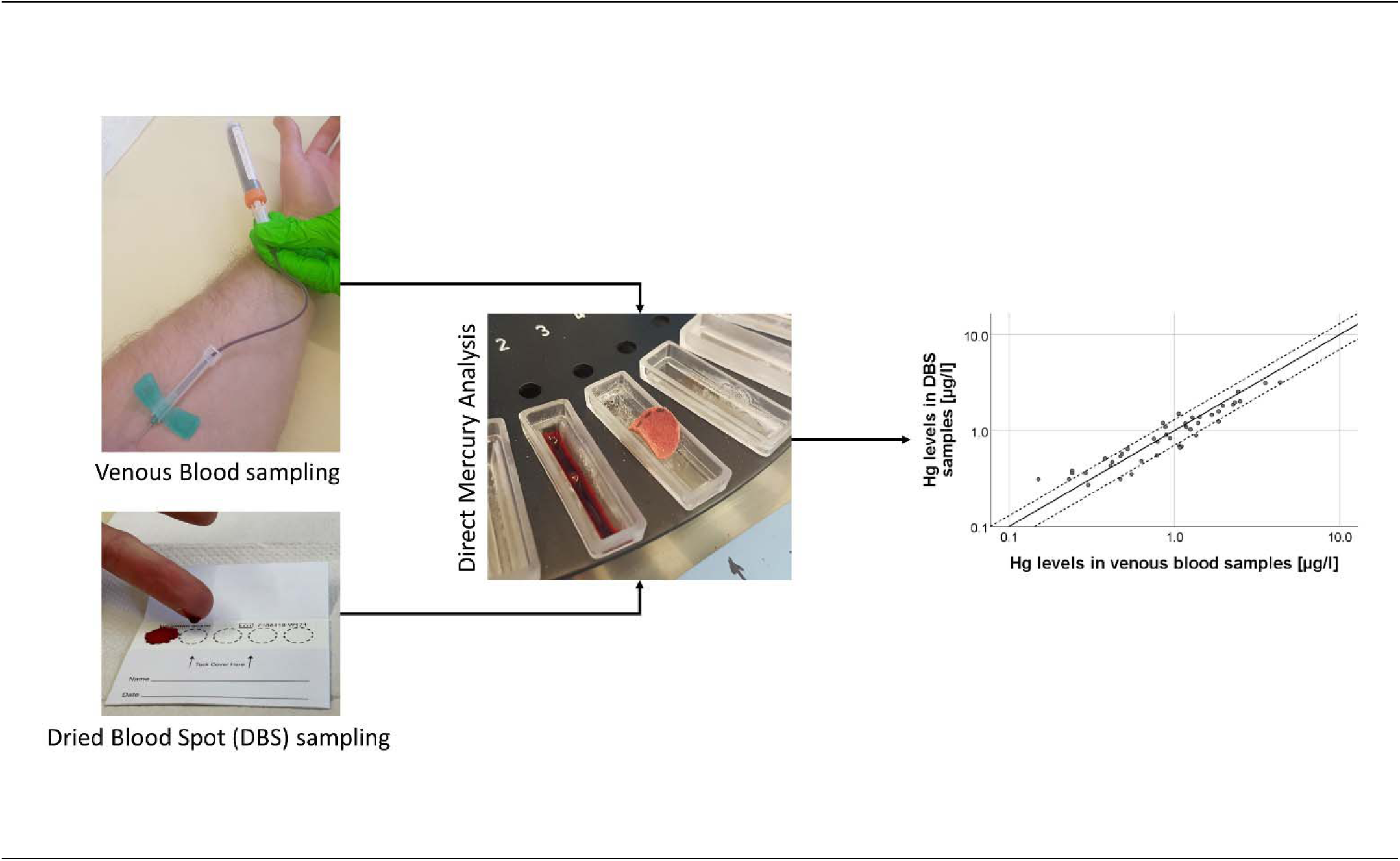

